# Assessment of prevalence and risk factors for intestinal parasitosis, malnutrition and anemia among school children in Ghindae area, Eritrea

**DOI:** 10.1101/2020.03.30.20042523

**Authors:** Yafet Kesete, Huruy Tesfahiwet, Ghimja Fessehaye, Yohana Kidane, Yafet Tekle, Asmerom Yacob, Biemnet Seltene

## Abstract

**Background:** Intestinal parasitic infections, anemia, and malnutrition are very endemic in resource-limited regions. School-aged children are at greater risk for the disease than any other age group as they are more susceptible to parasitosis, associated undergrowth and anemia. This study is aimed at evaluating the risk factors of intestinal parasitosis, malnutrition and anemia amongst elementary and junior school students in Ghindae area, Eritrea.

**Methods:** A cross sectional study was conducted in 6 schools around Ghindae from February to April 2018. 450 children were randomly selected for analysis and consent was taken from guardians. The pertinent sociodemographic data was collected and anthropometric measurements were carried out to determine the proportion of students with malnutrition, stunting and thinness. Fecal samples were examined by formal concentration technique and blood specimen was collected for the assessment of hemoglobin using hemocue analyzer.

**Results:** The overall prevalence of intestinal parasitosis was 45.3%. Protozoa infections (38.2%) were more prevalent than soil-transmitted helminthes infections (10.4%). The presence of different intestinal parasitic infections has statistically significant association with the residence area, washing habits, source of water, types of schools and type of latrine used with (p < 0.05). The prevalence of malnutrition was 37.1% with 18.5% stunting and 21.2% thinness. Malnutrition was in higher prevalence in semi-urban areas outside Ghindae. Students from Embatkala and Dongolo had 4.77 and 2.86 times higher odds of having low BMI for age than their counterpart respectively. The current prevalence of anemia was 12.4%, out of which, 7.6 % had mild while 4.4% of them had moderate anemia and 0.4% were severely anemic.

**Conclusion:** The prevalence of intestinal parasitic infections, especially, protozoan infection was very high in the school children. Stunting was increased rate in rural areas around Ghindae. The prevalence of anemia was regarded as mild health burden. The high prevalence of parasitic infections in these children indicates that the protozoa and helminthes concerned are very common in the environment of these area and results of the risk factors analysis suggest that the transmission is from several routes. Access to clean water and latrines, with some hygiene and sanitation communication activities, in addition to introduction of micronutrient programs could improve health of children in that area.

## Background

Intestinal parasitic infections, anemia, and malnutrition are highly prevalent in resource-limited countries. Intestinal helminthes are more prevalent throughout the tropics, especially among communities with low socio-economic level. Children are one of the groups at high-risk for intestinal parasitic infections [1]. In rural areas of developing countries, it is a major cause of morbidity in school age children, particularly in primary and junior school pupils (6 – 16 years) who harbor the greatest prevalence of worm infestation [2]. Distributions of IPIs are closely linked to lack of sanitation, shortage of access to safe water and proper hygiene practices; therefore, they are common whenever there is poverty.

Intestinal parasitosis is also an important conducing factor to poor nutritional and growth status and related child and infant mortality. Malnutrition during childhood also has a key significance for adult health and economic achievement [3]. Anemia’ refers to a condition in which the hemoglobin content of the blood is lower than normal as a result of deficiency of one or more essential nutrients, heavy blood loss, parasitic infections and congenital hemolytic diseases [4, 5]. Anemia is a very critical health issue because it influences the growth and energy levels of children and pregnant women adversely. Particularly children living in low income families are prone to develop iron deficiency anemia which occurs due to high demand of iron during period of fast growth [6].

The risk of a person suffering from IPI infection related morbidity is directly related to the joint function of the quantity of species harbored and/or the infection virulence or intensity of any species and are also associated with more risk for growth deficits and childhood malnutrition [7]. Intestinal parasitic infections and related malnutrition have detrimental effects on the survival, growth, appetite, physical fitness and school attendance of students [8]. Other considerable morbidities related to intestinal helminthes include anemia, growth retardation, vitamin A deficiency and have detrimental effect on students’ intellectual performance [8].

Parasitic infections like helminthiasis cause malnutrition [9] through different mechanisms including, increasing metabolic requirements, decreasing food intake and nutrient absorption up to direct loss of nutrient [10]. The relationship between intestinal parasitosis and anemia particularly microcytic hypochromic anemia and iron deficiency [11] is as well common and depends on the major parasites especially Hookworm infection, Schistosomiasis, and Amoebiasis.

To be effective, interventions aimed at reducing the effects of infection and malnutrition need to be based on a proper assessment of the current situation. No adequate previous studies have been conducted on the assessments of intestinal parasitosis, malnutrition, and anemia in Eritrea which could have been used as reference. Those which have been published are very few. Therefore, the study can be used as baseline data for better control and prevention strategies.

This study is aimed at evaluating the prevalence and risk factors of intestinal parasitosis, malnutrition and related anemia amongst elementary and junior school students in Ghindae area, northern Red Sea region, Eritrea. This study will identify the high risk population that is fundamental for appropriate resource allocation and cost effective control & allowing for reliable estimation of the overall drug-needs of programs and efficient geographical targeting of control efforts.

## Methods

### Study design and study population

Cross sectional study design was employed from February 2018 up to April 2018 in eastern Eritrea. Ghindae is located 48 km north-east of Asmara, the capital of Eritrea.The town along with several small towns is comprised in one administrative sub-zone in which around 12,000 families and approximately 54,000 people live within its boundary. The people’s life is mainly agricultural and some are involved in grazing and various commercial activities. At the altitude of 1020m, the Ghindae domain entertains three climates; cooler climate highlands, semi hot lowlands and lowland climate. Five years average annual rainfall for that place was 800 mm with bimodal rainy seasons. Most of inhabitants are subsistence farmers, cultivating their own plots of land for small-scale, rain-fed food production. There is one regional referral hospital along with few healthcare centers.

The total population of elementary and junior school students in Ghindae area is 7895 attending in 10 different schools with 3451 being females and 4444 being males. Using stratified random sampling technique 6 schools were selected from the list of schools in the area. 450 school children were selected from the preselected schools according to their proportion from the total number of Elementary and Junior school students. Students were selected using the class register as sampling frame. Students who have taken anthelminthic medication within two weeks before study and/or who were not able to submit both the samples were excluded from the study.

### Socio-demographic data collection

The pertinent data for the study was collected using a pre-tested, pre-coded questionnaire and was delivered to subjects interview-based to assess the baseline socio-demographic data as well as health status data, including previous or current medical and risk factors that are known to expose to intestinal parasitosis, malnutrition and anemia. The questionnaire was prepared for guardians and students individually. Most of the socio-demographic data was collected by interviewing the parents or guardians.

Observational exam was performed to study some variables, like hand hygiene and cleanliness of hand fingernails. Physical cleanliness of hands was evaluated by checking the finger pads, fingertips, palm, and the back of both hands. Hands were regarded as unclean if any noticeable dirt was seen and clean if there was no visible dirt. To assess finger nail hygiene of both hands, trimmed fingers were considered as clean and untrimmed nails with accumulated dirt were considered as unclean.

To ascertain a common understanding among all data collectors, discussion sessions and in house-practice programs were held using role play and thereafter pre-testing was conducted in the field prior to actual data collection on 42 subjects.

### Anthropometry

Anthropometric data like age, height and weight of the study participants was collected. A stadiometer with a sliding headpiece was used to record student’s height barefooted. Each student was also weighed with minimum clothing using a portable weight scale which was calibrated using standard calibration weights of 1 kg iron bars. The weight and height measurement of each participant was done twice and the average was recorded.

Anthropometric measurements were converted into height-for-age and body mass index (BMI)-for age Z scores using 2007 WHO growth reference [12]. Students which were below-2z score for weight-for age, body mass index-for age and height-for-age were regarded as underweight, thin, and stunted respectively.

### Stool specimen collection and analysis

Each participant was provided with a pre labelled, clean, wide mouthed 20ml stool container with screw caps instructing the gurdians to use the spatula attached to the cover of the bottle to collect fresh stool sample from their children the next morning. Subjects were advised on proper handling of stool samples & providing early morning faecal sample which usually present with more parasite than those collected at other time [13, 14]. All teachers were requested humbly in sample collection for full co-operation. About thumb size (10g) fresh stool specimens were collected from each subject. Each sample was fixed in 10% formalin immediately after collection and examined at Asmara College of Health Science parasitology laboratory within 24 hours after collection.

Stool samples were examined using standard procedure of formal-ether concentration technique [14]. From the emulsified sample 1 g (thumb size) of feces was added to about 4 ml of 10% formalin and then mixed and sieved in another tube. Then 3-4 ml of Diethyl ether was added and centrifuged immediately at 750-1000 g (∼ 3000 rpm) for 1 min. Finally the supernatant was discarded, and then small portion of the sediment was transferred to a slide and covered with cover slip and examined first with 10X and then 40X objectives and also iodine stained slides were prepared and examined microscopically.

### Blood specimen collection and analysis

Hemoglobin level of participants was determined using Hemocue analyzer measuring finger-prick blood sample in the school compound. A short training on the machine operation was given to research members before the actual sample collection period.

Hemoglobin level was divided into four for two age categories. For children 5 to 11 year; Above 11.5 g/dl normal, 11.0–11.4 g/dl mild anemia, 8.0–10.9 moderate, and < 8.0 g/dl severe anemia. For children 12 to 14 year; Above 12 g/dl normal, 11.0–11.9 g/dl mild anemia, 7.0– 10.9 moderate, and < 7.0 g/dl severe anemia. The machines were checked on a daily basis using the reference microcuvettes as indicated by the manufacturer. Hemoglobin readings were adjusted for altitude based on WHO standard [15].

### Statistical Analysis

The data was analysed using SPSS statistical software version 20. Descriptive statistics was used evaluate the data. Differences in the prevalence and intensity of infection between age and sex was tested using the Pearson Chi-square/ Fishers exacts test. Logistic Regression was used to evaluate the relative importance of specific categorical variables. One way ANOVA and independent t-tests was used to analyze hematological parameters at 5% significant level. Output data was presented using tables and figures.

## Results

From the 460 students selected for the research, 450 children were present with fully completed questionnaire and anthropometric measurement and were able to provide appropriate stool and blood specimens with a response rate of 97.6%. The study included school children aged 6-16 years out of which 215 (47.8%) were males and the mean (±SD) age was 10.34 (SD ± 2.6) years.

The result of the stool examination showed that 204 (45.3%), 95% CI: 40.9, 49.8 %) children tested positive for one or more intestinal parasitic infection (Table 1). Four intestinal parasite species i.e. *Entamoeba histolytica/dispar, Giardia lamblia, Hymenolepsis nana*, and Hookworm species were identified. The combination of *E. histolytica/dispar* and *G. lamblia* were the most predominant in double infection, accounting for (65.2%) of the total infected study subjects (Table 2).

**Table 1.** Prevalence of Different Parasites in Ghindae Area, Eritrea.

| Variables | Total Cases | Gender |  | Age |  | Residence |  |  |
| --- | --- | --- | --- | --- | --- | --- | --- | --- |
|  |  | Male(%) | Female(%) | 6-11(%) | 12-16 (%) | Ghindae(%) | Embatkala(%) | Dongolo(%) |
|  |  | 215(47.8) | 235(52.2) | 279(62) | 171(38) | 381(85.3) | 34(7.6) | 32(7.1) |
| Parasitosis | 204(45.3%) | 96 (47.1) | 108(52.9) | 131(64.2) | 73(35.8) | 181(47.3) | 13(32.4) | 20 (37.5) |
| Amoeba | 110(24.5%) | 52 (47.3) | 58 (52.7) | 61 (55.5) | 49(44.5) | 100(26.2) | 5(14.7) | 5 (15.6) |
| Giardia | 97(21.6%) | 42 (43.3) | 55 (56.7) | 70 (72.2) | 27(27.8) | 86 (22.5) | 6(17.6) | 5(15.6) |
| H.nana | 43(9.6%) | 21 (48.8) | 22 (51.2) | 33 (76.7) | 10(23.3) | 40 (10.5) | 2(5.5) | 3(9.4) |
| Hookworm | 5(1.1%) | 1(20) | 4(80) | 1(20) | 4(80) | 1(20) | 0(0) | 4(80) |
| Multiple infection | 46 (10.2%) | 16(34.8) | 30(65.2) | 29(63) | 17(37) | 42(23.2) | 0(0) | 4(33.3) |
| Malnutrition N (%) | 166 (37.1) | 82(38.1) | 84(35.7) | 89(31.9) | 77(45.0) | 131(34.2) | 19(55.9) | 16(50.0) |
| Stunting N (%) | 83(18.4) | 35(16.3) | 48(20.4) | 43(15.4) | 40(23.4) | 75 (19.6) | 2(5.9) | 6(18.8) |
| Low BMI for Age N(%) | 95(21.3) | 51(23.8) | 44(18.8) | 48(17.3) | 47(27.6) | 66(17.3) | 17(50.0) | 12(37.5) |
| Anemia | 56(12.4) | 20(9.3) | 36(15.3) | 30(10.8) | 26(15.2) | 48(12.5) | 1(2.9) | 7(21.9) |

**Table 2.**
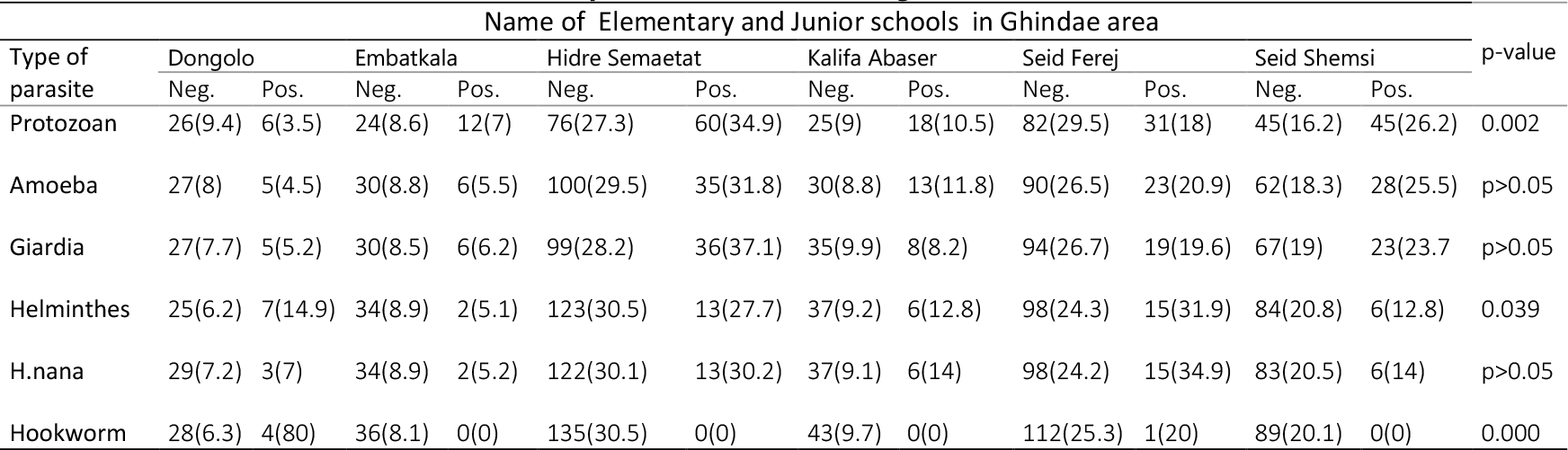
Distribution of instestinal infections among Ghindae area E.J. schools, Eritrea, 2018.

The overall prevalence of malnutrition was 37.1% (95% CI: 33.0, 41.7 %). Out of the studied schoolchildren, 18.5 %, were stunted (Z-score of HA < 2SD) and 21.2 % were wasted (Z-score of BMI < 2SD). Also severe stunting (HAZ <−3SD) and thinness (BAZ < −3SD) was observed with prevalence of 3.6% and 4.9% respectively. Prevalence of anemia among primary school children was 12.4 % (95% CI; 9.6, 15.6). The mean ± SD hemoglobin was 12.85 ± 1.19 g/dl. The mean ± SD value of hemoglobin in male children was 12.92 ± 1.26 g/dl and 12.78 ± 1.13 g/dl in female children. The anemic group was further classified into mild anemia which account for 7.6% and rest 4.4 % and 0.4% were moderately and severely anemic respectively. Children with age group 6–11 were more infected with *G. lamblia* and *H. nana* than 12–16 age group of study. Students who know the purpose of washing hands and with higher frequency of practice of hand washing had lower prevalence of parasitic infection. Similarly, poor hand hygiene was associated with higher infection percentages of *Giardia lamblia* cyst and *Hymenolepsis nana*. Students who had reported history of bloody diarrhea had higher prevalence of hookworm and *H. nana* infections. Also students who had been treated before for intestinal infection had Giardiasis significantly. Double infection was lower in prevalence in families who use their own latrine. Hookworm infection was found more in communities outside Ghindae town (Table 3). In addition, the Saho ethnic group had a statistically significant higher rate of protozoan infection compared to other ethnic groups. Students whose families use a common latrine along with other families had a significantly higher prevalence of Giardiasis and multiple infections. Using stream and river water as source for drinking was more exposing to protozoan infection (Table 3).

**Table 3.** Potential risk factors associated with Intestinal Parasites in Ghindae Area, Eritrea, 2018.

| Variables | Category | Total<br>N (%) | Protozoan |  |  | Helminthes |  |  |
| --- | --- | --- | --- | --- | --- | --- | --- | --- |
|  |  |  | N (%) | COR | p-value | N (%) | COR | p-value |
| **Gender** | Male | 215(47.8) | 80(37.2) | 1 | 0.67 | 22(10.2) | 1 | 0.88 |
|  | Female | 235(52.2) | 92(39.1) | 1.09 (0.74, 1.59) |  | 25(10.6) | 1.04 (0.570, 1.91) |  |
| **Age** | 6-11 | 279(62) | 108(38.7) | 1 | 0.95 | 34(12.2) | 1 | 0.12 |
|  | 12-16 | 171(38) | 64(37.4) | 0.95 (0.64, 1.40) |  | 13(7.6) | 0.59(0.30, 1.16) |  |
| **Ethnic Group** | Tigrigna | 156(34.7) | 50(32.1) | 1 | 0.13 | 22(14.1) | 1 | 0.24 |
|  | Tigre | 179(39.8) | 71(39.7) | 1.39(0.89, 2.19) | 0.15 | 14(7.8) | 0.52 (0.26, 1.05) | 0.068 |
|  | Saho | 110(24.4) | 50(45.5) | 1.77 (1.07, 2.93) | 0.03 | 10(9.1) | 0.61(0.28, 1.34) | 0.21 |
|  | Other | 5(1.1) | 1(20) | 0.530(0.058, 4.87) | 0.53 | 1(20) | 1.52 (0.16, 14.26) | 0.71 |
| **Maternal age** | 0-24 | 10(2.3) | 4(40) | 1 | 0.78 | 4(40) | 1 | 0.015 |
|  | 25-40 | 343(77.6) | 133(38.8) | 0.95 (0.26, 3.43) | 0.94 | 30(8.7) | 0.14 (0.04, 0.54) | 0.004 |
|  | >40 | 89(20.1) | 31(34.8) | 0.80 (0.21, 3.06) | 0.75 | 10(11.2) | 0.19 (0.05, 0.79) | 0.022 |
| **Occupation of mother** | Housewife | 354(80.1) | 138(39) | 1 | 0.62 | 38(10.7) | 1 | 0.11 |
|  | Merchant | 5(1.1) | 1(20) | 0.39(0.043, 3.54) | 0.40 | 2(40) | 5.54(0.898, 34.2) | 0.065 |
|  | Government worker | 38(8.6) | 12(31.6) | 0.72(0.35, 1.48) | 0.37 | 2(5.3) | 0.46 (0.11, 1.99) | 0.30 |
|  | Farmer | 2(0.5) | 0(0) | 0.00 | 0.99 | 1(50) | 8.31 (0.51, 135.6) | 0.14 |
|  | Other | 43(9.7) | 20(46.5) | 1.36(0.72, 2.57) | 0.34 | 3(7) | 0.62 (0.18, 2.11) | 0.45 |
| **Hand Hygiene** | Clean | 286(63.3) | 78(47.6) | 1 | 0.002 | 24(14) | 1 | 0.06 |
|  | Unclean | 164(36.4) | 94(32.9) | 1.85 (1.25, 2.75) |  | 23(8.4) | 1.78 (0.97, 3.27) |  |
| **Knowing Purpose of washing Hands** | Yes | 129(28.7) | 152(36.5) | 1 | 0.012 | 43(10.3) | 1 | 0.79 |
|  | No | 34(7.6) | 20(58.8) | 2.48(1.21,5.05) |  | 4 (11.8) | 1.16 (0.39,3.44) |  |
| **Residence area** | Ghindae | 383(85.3) | 155(40.5) | 1 | 0.048 | 40(10.4) | 1 | 0.56 |
|  | Embatkala | 34(7.6) | 11(32.4) | 0.70(0.33,1.49) | 0.35 | 5(9.1) | 0.90(0.33,1.49) | 0.46 |
|  | Dongolo | 32(7.1) | 6(18.8) | 0.34 (0.137,0.84) | 0.020 | 7(21.9) | 2.04 (0.98, 5.90 ) | 0.33 |
| **Water Source** | River/Spring | 86(19.1) | 37(43) | 1 | 0.036 | 8(9.3) | 1 | 0.014 |
|  | Pipe | 323(71.8) | 127(39.3) | 0.86(0.53, 1.40) | 0.54 | 29(9) | 0.96(0.42, 2.19) | 0.96 |
|  | Water truck | 41(9.1) | 8(19.5) | 0.32 (0.13, 0.78) | 0.012 | 10(24.4) | 3.15 (1.14, 8.71) | 0.03 |
| **History of Bloody diarrhea** | No | 321(89.4) | 128(39.9) | 1 | 0.71 | 27(8.4) | 1 | 0.017 |
|  | Yes | 38(10.6) | 14(36.8) | 0.89 (0.44, 1.76) |  | 8(21.1) | 2.91 (1.21, 6.96) |  |
| **Treatment in last month** | No | 252(70.8) | 92(36.5) | 1 | 0.09 | 26(10.3) | 1 | 0.84 |
|  | Yes | 104(29.2) | 48(46.2) | 1.49(0.94, 2.37) |  | 10(9.6) | 0.93 (0.43, 1.99) |  |
| **Place of defecation at school** | Latrine | 212(47.1) | 73(34.4) | 1 | 0.119 | 22(10.4) | 1 | 0.96 |
|  | Other | 238(52.9) | 99(41.6) | 1.36 (0.93, 1.99) |  | 25(10.5) | 1.01 (0.55, 1.86) |  |
| **Stool consistency** | Formed | 183(40.7) | 54(29.5) | 1 | 0.012 | 18(9.8) | 1 | 0.98 |
|  | Soft | 219(48.7) | 94(42.9) | 1.80 (1.19, 2.72) | 0.006 | 24(11) | 1.13( 0.59, 2.15) | 0.71 |
|  | Loose | 45(10) | 23(51.1) | 2.50 (1.28, 4.86) | 0.007 | 5(11.1) | 1.12 (0.40, 3.27) | 0.79 |
|  | Watery | 3(0.7) | 1(33.3) | 1.19 (0.11, 13.5) | 0.886 | 1(30) | 1.13(0.75, 1.72) | 0.99 |

In bivariate logistic regression, age, residence area, type of school, grade and waste disposal were found to be significantly associated with malnutrition. Students aged 12-16 had 1.68 times higher odds of stunting, 1.75 times greater odds of malnutrition and 2.23 times higher odds of having low BMI for age compared to those aged 6 to 11. Thinness and malnutrition was in higher prevalence in semi-urban areas outside Ghindae. Peripheral areas like Embatkala and Dongolo area residents had 4.77 and 2.86 times more odds of having low BMI for age than their counterparts.

The prevalence of malnutrition was not affected by parental educational status, ethnicity and religion. There was no statistically significant association between anemia and thinness. Female students had higher prevalence of moderate anemia than their counterparts. Higher prevalence of hookworm infection was present in anemic students (40%), compared with 12.2% of anemic school children who had no hookworm (Table 4).

**Table 4.** Potential risk factors associated with Malnutrition, Stunting, Underweight and Anemia in Ghindae Area, Eritrea, 2018.

|  | Category | Malnutrition |  |  |  | Low BMI for age |  |  | Low Height for age |  |  | Anemia |  |  |
| --- | --- | --- | --- | --- | --- | --- | --- | --- | --- | --- | --- | --- | --- | --- |
|  |  | Total<br>N (%) | N (%) | COR | p-value | N (%) | COR | p-value | N (%) | COR | p-value | N (%) | COR | p-value |
| **Gender** | Male | 215(47.8) | 82(38.1) | 1 | 0.59 | 51(23.8) | 1 | 0.19 | 35(16.3) | 1 | 0.26 | 20(9.3) | 1 | 0.053 |
|  | Female | 235(52.2) | 84(35.7) | 0.90 (0.62, 1.32) |  | 44(18.8) | 0.74(0.47,1.17) |  | 48(20.4) | 1.32(.82,2.14) |  | 36(15.3) | 1.77 (0.98,3.15) |  |
| **Age** | 6-11 | 279(62) | 89(31.9) | 1 | 0.005 | 48(17.3) | 1 | 0.01 | 43(15.4) | 1 | 0.035 | 30(10.8) | 1 | 0.056 |
|  | 12-14 | 171(38) | 77(45.0) | 1.75(1.18,2.59) |  | 47(27.6) | 1.83(1.16,2.89) |  | 40(23.4) | 1.68(1.04, 2.71) |  | 26(15.2) | 1.48(0.85,2.62) |  |
| **School name** | Dongolo | 32(7.1) | 16(50) | 1 | 0.002 | 12(37.5) | 1 | 0.001 | 6(18.8) | 1 | 0.001 | 7 (21.9) | 1 | 0.12 |
|  | Embatkala | 36(8) | 20(55.6) | 1.25(0.48,3.25) | 0.65 | 18(50) | 1.66(0.63,4.39) | 0.30 | 2(5.6) | 0.26(0.05 1.37) | 0.11 | 1(2.8) | 2.87 (0.95,8.69) | 0.06 |
|  | Hidre-Semaetat | 136(30.2) | 46(33.8) | 0.51(0.24, 1.11) | 0.09 | 20(14.7) | 0.29(0.12,0.68) | 0.004 | 29(21.3) | 1.17(0.44, 3.12) | 0.75 | 15(11) | 0.29 (0.04,2.43) | 0.26 |
|  | Khalifa | 43(9.6) | 5(11.6) | 0.13(0.041,0.42) | 0.001 | 5(11.6) | 0.22(0.07,0.71) | 0.011 | 1(2.3) | 0.10(0.01,0.91) | 0.04 | 5(11.6) | 1.27 (0.51,3.13) | 0.60 |
|  | Seid Ferej | 113(25.1) | 41(36.3) | 0.57(0.26,1.26) | 0.16 | 30(26.8) | 0.61(0.26,1.39) | 0.24 | 15(13.3) | 0.66(0.23,1.88) | 0.44 | 20(17.7) | 1.34 (0.41,4.39) | 0.62 |
|  | Seid Shemsi | 90(20.0) | 38(42.2) | 0.73(0.33,1.64) | 0.45 | 10(11.2) | 0.21(0.08,0.56) | 0.002 | 30(33.3) | 2.17(0.81,5.83) | 0.13 | 8(8.9) | 2.20 (0.92,5.27) | 0.07 |
| **Grade** | 1-3 | 174(38.7) | 55(31.6) | 1 | 0.014 | 25(14.5) | 1 | 0.004 | 32(18.4) | 1 | 0.34 | 23(13.2) | 1 | 0.62 |
|  | 4-5 | 112(24.9) | 36(32.1) | 1.03(0.62,(1.71) | 0.93 | 22(19.6) | 1.45(0.77,2.72) | 0.25 | 16(14.3) | 0.74(0.38,1.42) | 0.36 | 11(9.8) | 0.98(0.52,1.84) | 0.96 |
|  | 6-8 | 164(36.4) | 75(45.7) | 1.82(1.17,2.84) | 0.008 | 48(29.4) | 2.47(1.44,4.25) | 0.001 | 35(21.3) | 1.20(0.71,2.05) | 0.49 | 22(13.4) | 0.70(0.32,1.51) | 0.37 |
| **Mother Occupation** | Housewife | 354 (80.1) | 123(34.7) | 1 | 0.06 | 69(19.5) | 1 | 0.12 | 65(18.4) | 1 | 0.84 | 49 | 1 | 0.07 |
|  | Other | 88(19.9) | 40(45.5) | 1.57(0.98, 2.51) |  | 24(27.6) | 1.57(0.92,2.69) |  | 17(19.3) | 1.07(0.59,1.93) |  | 6 | 0.46 (0.19, 1.10) |  |
| **Residence area** | Ghindae | 383(85.3) | 131(34.2) | 1 | 0.014 | 66(17.3) | 1 | 0.000 | 75(19.6) | 1 | 0.18 | 48(12.5) | 1 | 0.09 |
|  | Embatkala | 34(7.6) | 19(55.9) | 2.44(1.20, 4.95) | 0.014 | 17(50) | 4.77 (2.32,9.83) | 0.000 | 2(5.9) | 0.26 (0.06,1.10) | 0.06 | 1(2.9) | 0.211(0.28, 1.58) | 0.13 |
|  | Dongolo | 32(7.1) | 16(50) | 1.93(0.94, 3.97) | 0.07 | 12(37.5) | 2.86(1.34,6.14) | 0.007 | 6(18.8) | 0.95 (0.37,2.39) | 0.91 | 7(21.9) | 1.95 (0.80, 4.76) | 0.14 |
| **Waste disposal** | Near home | 106(23.6) | 49(46.2) | 1 | 0.023 | 31(29.2) | 1 | 0.02 | 21(19.8) | 1 | 0.67 | 12(11.3) | 1 | 0.68 |
|  | Far from home | 344(76.4) | 117(34) | 0.6(0.38,0.93) |  | 64(18.7) | 0.55(0.33,0.91) |  | 62(18.0) | 0.89(0.51,1.54) |  | 44(12.8) | 0.87(0.44,1.71) |  |
| **Parental Status** | Living Together | 289(65.4) | 109(37.7) | 1 | 0.059 | 62(21.5) | 1 | 0.104 | 54(18.7) | 1 | 0.45 | 37(12.8) | 1 | 0.60 |
|  | Divorce | 27(6.1) | 14(51.9) | 1.77 (0.81,3.92) | 0.154 | 10(37.0) | 2.15(0.94,4.94) | 0.07 | 4(14.8) | 0.76(0.25,2.28) | 0.62 | 2(7.4) | 0.89(0.46,1.7) | 0.72 |
|  | Widowed | 20(4.5) | 2(10) | 0.18(0.04,0.81) | 0.25 | 1(5) | 0.19(0.03,1.47) | 0.11 | 1(5.0) | 0.23(0.03,1.75) | 0.16 | 1(5.0) | 0.48(0.10,2.26) | 0.35 |
|  | Separated | 106(24) | 39(36.8) | 0.96(0.61, 1.52) | 0.87 | 22(21.2) | 0.98(0.57,1.70) | 0.95 | 22(20.8) | 1.14(0.65,1.98) | 0.64 | 15(14.2) | 0.32(0.04,2.56) | 0.28 |

## Discussion

The study showed widespread prevalence of intestinal parasitosis among school children in Ghindae area. The overall prevalence of intestinal parasite was (45.3%) was consistent with the studies conducted at different parts of Africa [16, 17, 18]. Results of study also showed that protozoan infections (38.2%) were more common compared with STH infections (10.4%) unlike other studies which report the opposite [19]. Double infection that is mainly due to Amoebiasis and Giardiasis was higher in Ghindae town probably attributed to the drinking water source that is mainly from nearby local river crossing through town on which almost all of households dispose their household and human wastes. Moreover, the climate is warm and with high humidity which directly relate to high consumption of water among residents making it favorable for protozoan infection which is mainly water borne disease. The hot and humid weather and wet soil may have favored for the presence of the parasite in this study area. Prevalence of *E. Histolytica/ dispar* (24.3%), the most frequently found parasite, along with *G. lamblia* (21.6%) was higher compared to other studies conducted elsewhere [20, 21]. *H. nana* had a prevalence rate 9.6% showing poor hygienic practices and was comparable to the report from the study conducted in Ethiopia [22] but it higher than the findings of other studies [23].

The Saho ethnic group had statistically significant higher rate of protozoan infection probably attributed to isolated living style in rural vicinity in which sole source of water is the river. In consistence with other studies, students who don’t know the purpose of washing hands and with dirt in their fingernails had 2.72 and 1.99 more likelihood of having parasitic infection [24, 25]. Compared to subjects without usage of latrines, lower prevalence rates of overall and *H. nana* infections among private or public latrine users was seen which was in line with other studies [26]. Students whose family water supply from river-stream were 3.12 more likely to have protozoan infection which is more associated with unsafe water sources.

Most of the schools don’t have functioning toilets contributing to the problem by practice of open field defecation at the nearby river. Trade and economic activities is more at the center of town and students are exposed to street food which upon sharing can be the main reason for the transmission dynamics. Hookworm was significantly prevalent in Dongolo area attributed to the rural and subsistence farming lifestyle receiving bimodal rainfall which is conducive for maturation of hookworm filariform larvae [27].

The overall prevalence of malnutrition in this study was 37.1%. and the most frequent type of malnutrition was low BMI for age (21.3%) which was within the range reported for Africa [28] and in agreement with other studies [29]. High stunting could be due to a prolonged shortage of balanced meals, especially amongst children from poor families [30]. The proportion of malnourished children was higher in the children aged 12-16 than that of 6-11 consistent with other study in Ethiopia [31].

Thinness and malnutrition was in higher prevalence in semi urban areas outside Ghindae in agreement with other studies [32]. Students living in Embatkala and Dongolo, semi-urban areas around the town, had 4.77 and 2.86 times higher odds of having low BMI for age than their counterpart respectively. This may be attributed to the socio-economic environment as well as variation in rural and urban settings in Eritrea. Similar to other studies, students whose mothers’ occupation is other than housewife had higher prevalence of malnutrition [29] because students will be overlooked after their dietary intake when their mothers spend their time away. Also students who have parents currently divorced had higher proportion of low BMI for age (thinness) on account of low socioeconomic status and income present in students with divorced parents.

In sharp contrary to some studies [33], but in agreement with previous study done in Ethiopia [34], anthropometric scores were not associated with overall rate of intestinal parasitic infections(p>0.05). The lack of association may be explained by the very low parasitic load or could be due to no measurable differences.

The magnitude of anemia determined in this study (12.4%) is considered a mild public health problem according to WHO standards [5]. The prevalence rates of mild and moderate and severe (Hb < 7 g/dl) anemia were 7.6%, 4.4 %, 0.4% respectively. Anemia prevalence in the study group was consistent with a recent prevalence report from neighboring countries [25] but lower than other studies in around the world [6, 35, 36] as determined by the same techniques.

Adolescents (12-16 years) had higher prevalence of anemia (15.2%) than their counterpart in agreement with previous studies [37]. Maternal education did not show significant association with anemia in contrary to findings of several studies [38, 39] probably due to the homogeneity of the respondents’ educational status as those who had no formal education were in the education for elders’ program which targets for elimination of illiteracy.

Similar to other studies, increased number of family members was significantly associated with severity of anemia (p=0.001) [40]. Unlike other studies, Income of families was not observed to be related to anemia [35, 40] as estimate of family income was derived from salary house hold head that doesn’t include other sources of income like remittance income present in significant part of sample size and inability to assess unofficial jobs like day to day work.

## Conclusion

This study demonstrated the effect of different socio-demographic factors and determinants on the parasitic infection, malnutrition and anemia. Though there might be many factors for the significantly increased protozoan infection in that area especially in periphery of town, unsafe drinking water sources from streams and springs are among the core problems. Practice of using human fertilizer observed in Dongolo areas and inappropriate waste disposal system seen in Ghindae town might have contributed to the burden in addition to absence of functioning toilets that can be accessed by students at home and school.

The magnitude of anemia determined in this study is considered as a mild public health problem as WHO recommends supplementation for women of childbearing age including adolescent girls and school-aged children, if anemia prevalence exceeds 40%. This study is beneficial exploratory reference study on risk factors leading to intestinal parasitosis malnutrition and anemia in the study area. Emphasis on the need of increased personal hygiene practice; such as increased access to clean water for hand washing and drinking, access to latrines to restrict contamination, and hygiene-related health education would improve students’ health. The cross-sectional nature of the study, made any inference on causal relationship among variables impossible.

## Data Availability

The data used to support the findings of this study are available from the corresponding author upon request.

## Abbreviations used

IPI: Intestinal parasitic infection
WHO: world health orqanization
Hb: hemoqlobin

## Ethics approval and consent to participate

Ethical clearance was acquired from the ACHS research ethical committee and Ministry of Health (MOH). The researchers obtained verbal and written consent from guardians of student on the consent form attached with questionnaire, and it remained anonymous. Capillary blood collection was performed after obtaining a signed written informed consent from parents and an oral assent from participant. Only student code number was used to retrieve the parasitological, anthropometric & hemoglobin results and were held confidential. Report of positive individuals was notified to concerned parties for proper treatment according to standard guidelines.

## Consent for publication

Not applicable.

## Competing interests

The authors declare that they have no competing interests.

## Funding

The study was funded partially by Asmara Collage of Health Sciences.

## Authors’ contributions

Y.K was the primary researcher, conceived the study, designed, participated in data collection and specimen analysis, conducted demographic and laboratory data analysis and drafted the manuscript for publication. H.T and Y.T participated in research study design and coordination and specimen analysis. G.F participated in conception of study, research study design and coordination and revised the final manuscript. B.S, Y.K and A.Y assisted in data collection and blood and stool specimen analysis and preparation of the first draft of the manuscript. All authors read and approved the final manuscript.

## Acknowledgment

This study was conducted with support of Asmara college of Health science and Ghindae Hospital. We are also thankful to all school children and their families who participated in the study and many people that assisted in this study. We wish to acknowledge the cooperation of the Ghindae subzone Administration, Education office of Ghindae, Halibet laboratory, National blood bank, Ghindae School of Nurse Assistants.

## Additional files

**Additional file 1:** Questionnaire used for the Assessment of prevalence and risk factors for intestinal parasitosis, malnutrition and anemia among school children in Ghindae area, Eritrea

## Notes

### Competing Interest Statement

The authors have declared no competing interest.

### Funding Statement

This study was funded partially by Asmara College of Health Sciences. Lab equipments required for stool and blood sample analysis was acquired from the college.

